# The impact of the coronavirus disease 2019 (COVID-19) outbreak on hospital admissions for alcohol-related liver disease and pancreatitis in Japan: a time-series analysis

**DOI:** 10.1101/2020.11.16.20232181

**Authors:** Hisashi Itoshima, Jung-ho Shin, Daisuke Takada, Tetsuji Morishita, Susumu Kunisawa, Yuichi Imanaka

## Abstract

**Background:** During the coronavirus disease 2019 (COVID-19) pandemic, there have been health concerns related to alcohol use and misuse. Therefore, the World Health Organization cautioned that alcohol consumption during the pandemic might have a negative impact. The aim of this study was to examine the population-level change in cases of alcohol-related liver disease and pancreatitis that required admission during the COVID-19 outbreak.

**Methods:** We included patients aged 18 years or older who were hospitalized between July 2018 and June 2020 using Diagnostic Procedure Combination data, an administrative database in Japan, and counted the admission cases whose primary diagnosis was alcohol-related liver disease or pancreatitis. We defined the period from April 2020, when the Japanese government declared a state of emergency, as the beginning of the COVID-19 outbreak. The rate ratio (RR) of admissions with alcohol-related liver disease or pancreatitis per 1,000 admissions was tested using interrupted time series analysis. In addition, excess admissions for alcohol-related liver disease or pancreatitis were calculated.

**Results:** Overall admissions were 3,026,389 cases, and a total of 10,242 admissions for alcohol-related liver disease or pancreatitis occurred from 257 hospitals. The rate of admissions per 1,000 admissions during the COVID-19 outbreak period (April 2020 to June 2020) had a 1.2 times increase compared with the pre-outbreak period (July 2018 to March 2020) for cases of alcohol-related liver disease or pancreatitis (RR: 1.22, 95%Confidence interval [CI]: 1.12 to 1.33). The COVID-19 pandemic caused about 214.75 (95%CI: 178.78 to 249.72) excess admissions for alcohol-related liver disease or pancreatitis based on predictions from our model.

**Conclusion:** The COVID-19 outbreak might have resulted in increased hospital admissions for alcohol-related liver disease or pancreatitis.

## INTRODUCTION

Alcohol misuse is a major public health concern that causes about 3 million deaths worldwide each year [1]. During the coronavirus disease 2019 (COVID-19) pandemic, there have been health concerns related to alcohol use and misuse. Therefore, the World Health Organization cautioned that alcohol consumption during the pandemic might have a negative impact such as risk-taking behaviors, mental health problems and violence [2]. Stress is a risk factor for alcohol misuse. Policies such as keeping social distance and isolation could cause people stress. Therefore, it is recommended that governments give warnings about excessive alcohol consumption during isolation [3].

Actually, since the stay-at-home policy began in some US states, one company has seen a 54% increase in national sales of alcohol per week, compared with 1 year before [4]. Some studies from the US and UK have reported an increased volume of alcohol consumption in households [5, 6]. In Japan, a survey from the Japanese government reported that the expenditure in households for alcohol after April 2020 increased by 40–50% compared with 1 year before [7].

Alcohol misuse also causes physical illness such as liver disorder and pancreatitis, alcohol-attributable fractions (AAAFs) for all global death, Disability-adjusted life years were accounted for 48%, 49% in liver cirrhosis and 26%, 28% in pancreatitis, respectively [1]. However, there have been few studies related to alcohol-related liver disease and pancreatitis during the COVID-19 pandemic.

This study examined the population-level change in cases of alcohol-related liver disease and pancreatitis that required admission during the COVID-19 outbreak.

## METHODS

### Study design

We conducted a quasi-experimental, interrupted time series analysis using Diagnosis Procedure Combination (DPC) data from the Quality Indicator/Improvement Project (QIP) database, which is administered by the Department of Healthcare Economics and Quality Management, Kyoto University. The QIP database consists of DPC data from acute care hospitals voluntarily participating in the project. The participating hospitals exceed 500, which are located throughout Japan and contain both public and private hospitals [8].

In Japan, the DPC/pre-diem payment system (PDPS) is a prospective payment system that is accepted for acute care hospitals. The number of general beds adopted by the DPC/PDPS in 2018 accounted for 54% of total general beds in Japanese hospitals (482,618/891,872) [9, 10]. The DPC data include insurance claims and clinical summary data, which contains facility identifiers, admission and discharge statuses (in-hospital death or alive discharge), cause of admission, primary diagnosis, the most and second -most medical resource-intensive diagnoses, up to 10 comorbidities and 10 complications. All diagnoses were classified according to the International Classification of Diseases, 10th Revision (ICD-10) codes. A published paper described further details [7].

### Data collection

We included patients aged 18 years or older who were hospitalized between July 1, 2018 and June 30, 2020 from the database, and counted the admission cases whose both primary and most-medical resource-intensive diagnoses were alcohol-related liver disease or pancreatitis based on ICD-10 codes (K70.1: Alcoholic hepatitis, K70.2: Alcoholic fibrosis and sclerosis of the liver, K70.3: Alcoholic cirrhosis of liver, K70.4: Alcoholic hepatic failure, K70.9: Alcoholic liver disease, unspecified, K85.2: Alcohol-induced acute pancreatitis, K86.0: Alcohol-induced chronic pancreatitis). We defined K70.1, K70.2, K70.3, K70.4 and K70.7 as alcohol-related liver disease and K85.2, K86.0 as alcohol-related pancreatitis.

### Outcomes of interest

The main outcome was the rate ratio (RR) of hospital admissions with alcohol-related liver disease or pancreatitis per 1,000 hospital admissions. Secondary outcomes were the RR of hospital admissions with alcohol-related liver disease, liver cirrhosis, acute pancreatitis and chronic pancreatitis per 1,000 hospital admissions, respectively. In addition, excess hospital admissions for alcohol-related liver disease or pancreatitis were calculated.

### Statistical analyses

To compare year-on-year, we divided the study population into two: those who were admitted and discharged from July 2018 to June 2019 and from July 2019 to June 2020. Because the DPC data were generated on the day of discharge, no data were available for patients who had not been discharged by the end of June 2020, even if their admission date was before June 30, 2020.

We conducted an interrupted time series (ITS) analysis using segmented and Poisson regressions by considering seasonality, trends and overdispersion of data, to analyze the outcomes [11–13]. Seasonality was taken into consideration by adding harmonic terms (sines and cosines) with 12-month periods to our model [11]. The validity of the Poisson regression model was evaluated by using the correlograms (functions of autocorrelation and partial autocorrelation) and the residuals. Excess hospital admissions for alcohol-related liver disease or pancreatitis was defined as the difference in hospital admissions between the actual number of hospital admissions and the predicted number of admissions based on our model.

We defined the period from April 2020, when the Japanese government declared a state of emergency, as the beginning of the COVID-19 outbreak, and assumed that the COVID-19 outbreak rapidly affected the level of hospitalization after April 2020 [14]. Several studies have found different alcohol consumptions by age (among younger and older adults) and sex [5, 15]. Therefore, we conducted an exploratory stratified analysis for sex and cases aged below and above 65 years.

A two-sided P-value <0.05 was considered statistically significant, and all analyses were performed using R 3.6.3 (R Foundation for Statistical Computing, Vienna, Austria).

### Ethical considerations

The present study was approved by the ethics committee of Kyoto University (approval number: R0135), and we did not require informed consent because of the use of anonymized data, in accordance with the Ethical Guidelines for Medical and Health Research Involving Human Subjects, as stipulated by the Japanese Government. There were no conflicts of interest in the manuscript.

## RESULTS

Overall hospital admissions were 3,026,389 cases, and 10,242 hospital admissions for alcohol-related liver disease or pancreatitis occurred in 257 hospitals, which had a total of 67,609 general beds during the study period.

The characteristics of the study population such as age, sex, length of hospital stay and in-hospital mortality did not change significantly between the pre-COVID-19 outbreak period (July 2018 to March 2020) and the COVID-19 outbreak period (April 2020 to June 2020) (Supplement Table 1). The monthly number of hospital admissions with alcohol-related liver disease or pancreatitis, a year-on-year comparison and the monthly rates per 1,000 hospital admissions are shown in Table 1. Figure 1 displays these data together with the predicted regression curves. We observed an increase in the rate of alcohol-related liver disease and pancreatitis immediately after the declaration of emergency for the COVID-19 pandemic by the Japanese government.

**Table 1.**
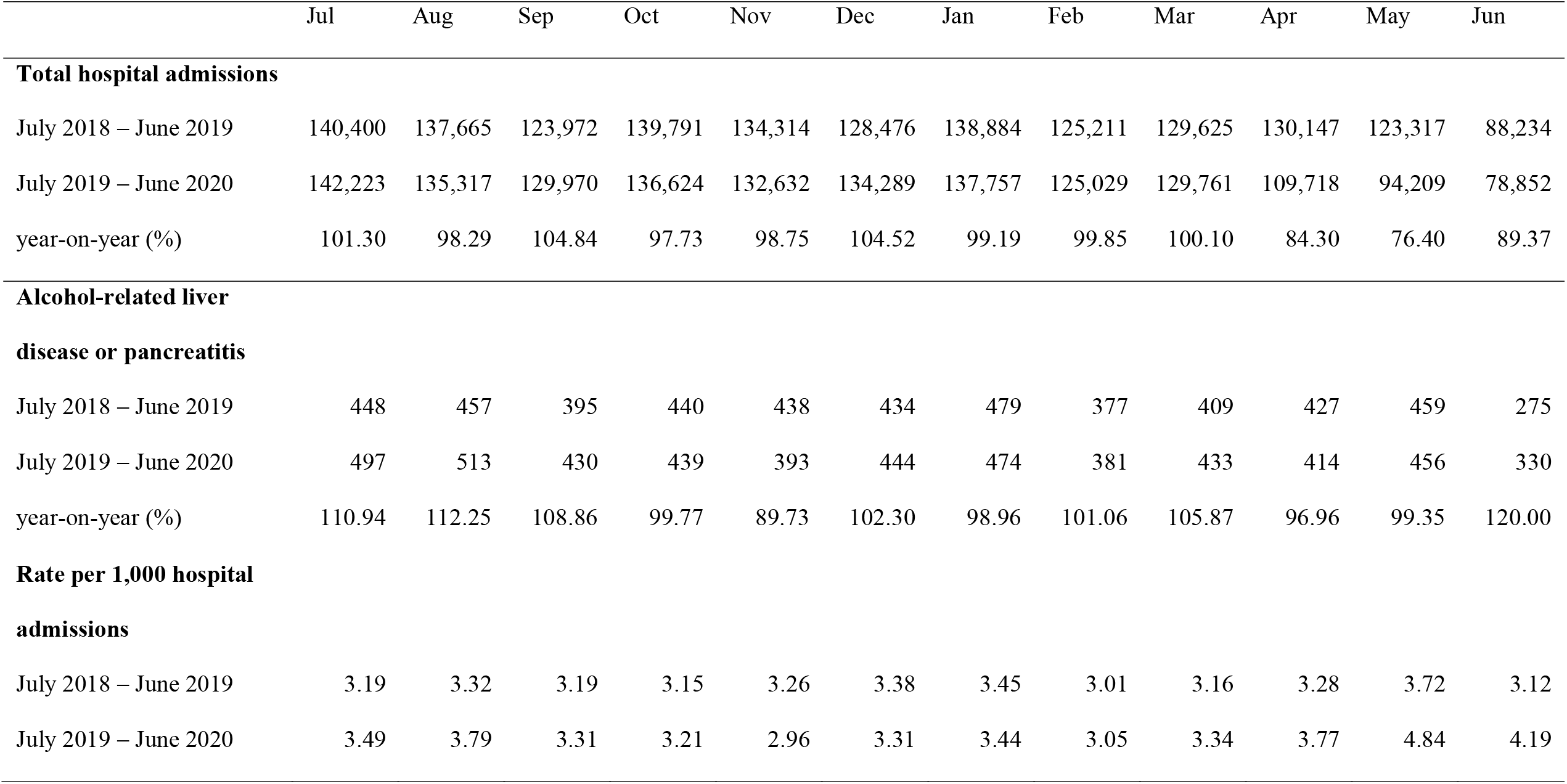
Number of hospital admissions for alcohol-related liver disease or pancreatitis, and rates (cases/1,000)

**Figure 1.**
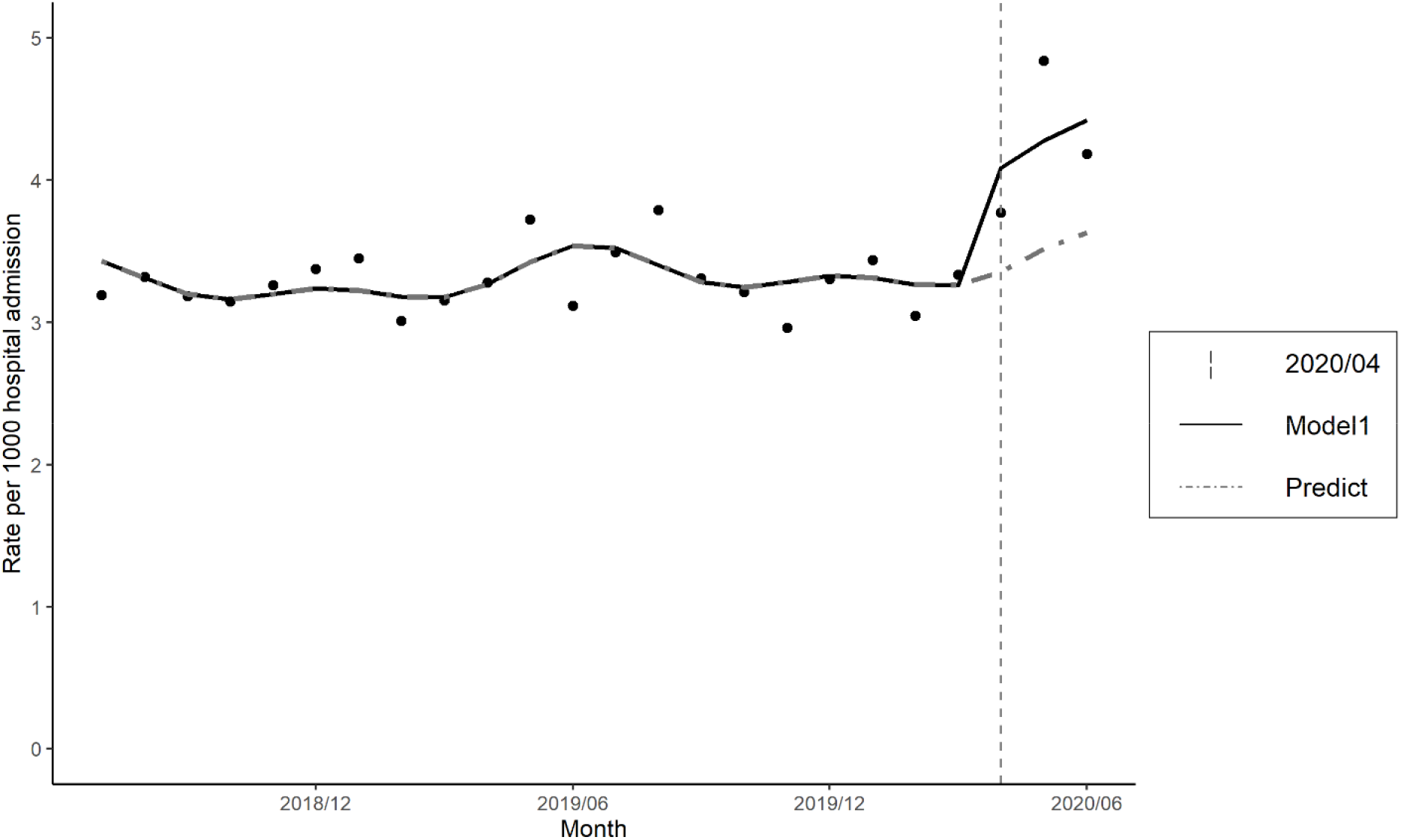
Hospital admissions for alcohol-related liver disease or pancreatitis during the study period. Model1: Poisson regression model, including trends and seasonality Predict: Counterfactual scenario (if the outbreak did not occur)

The rate ratio (RR) for alcohol-related liver disease or pancreatitis during the COVID-19 outbreak period (April 2020 to June 2020) compared with the pre-COVID-19 outbreak period (July 2018 to March 2020) was 1.22 (95% confidence interval [95%CI] 1.12 to 1.33). Under the counterfactual scenario (if the COVID-19 outbreak did not occur), our model predicted 985.75 (95%CI: 968.38 to 1,003.12) hospital admissions for alcohol-related liver disease or pancreatitis from April 2020 to June 2020, whereas 1,200 actual hospital admissions occurred during these three months. Therefore, this indicated that 214.25 (95%CI: 196.88 to 231.62) excess hospital admissions took place during the COVID-19 outbreak period (Supplement Table 2).

**Table 2.**
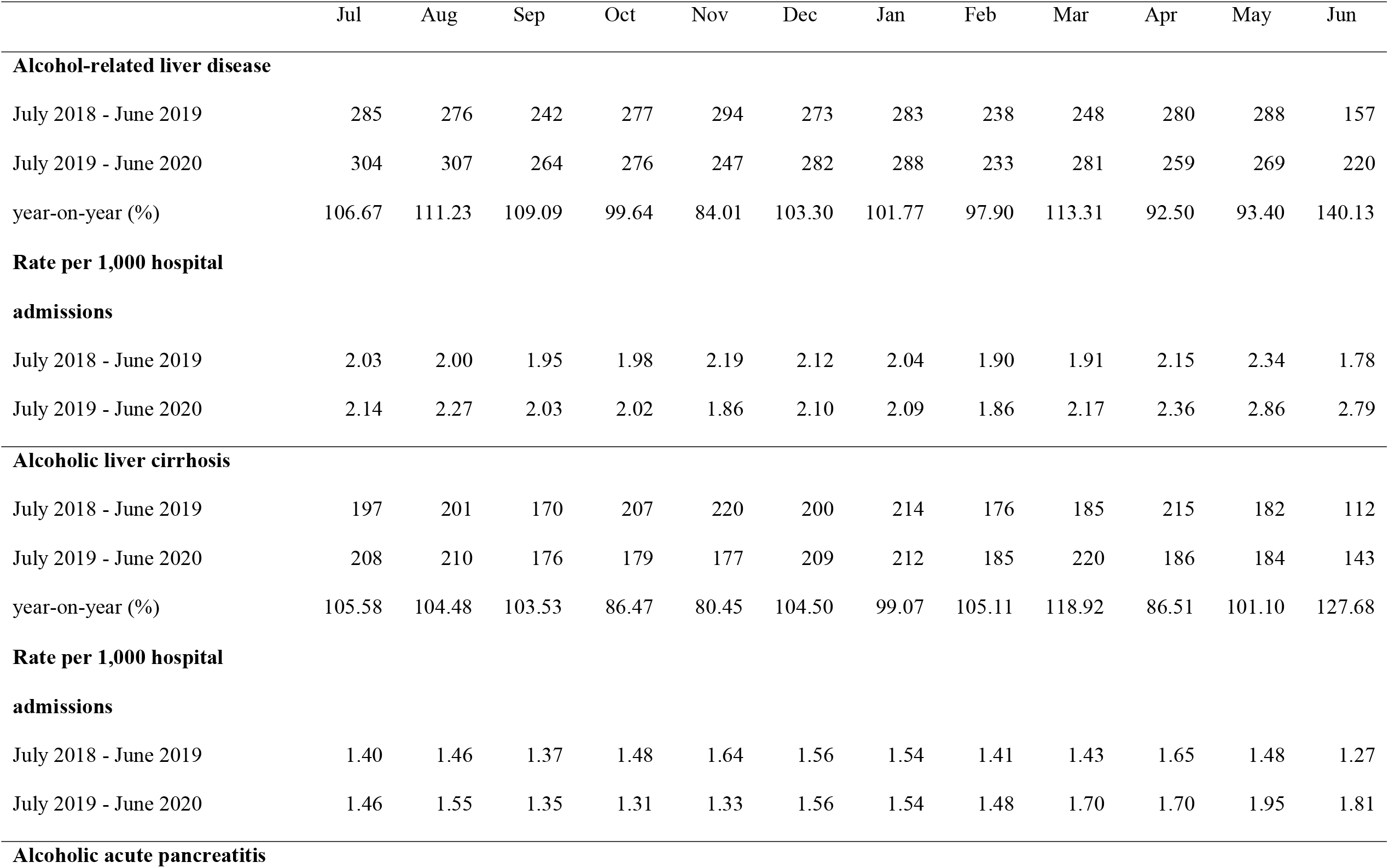
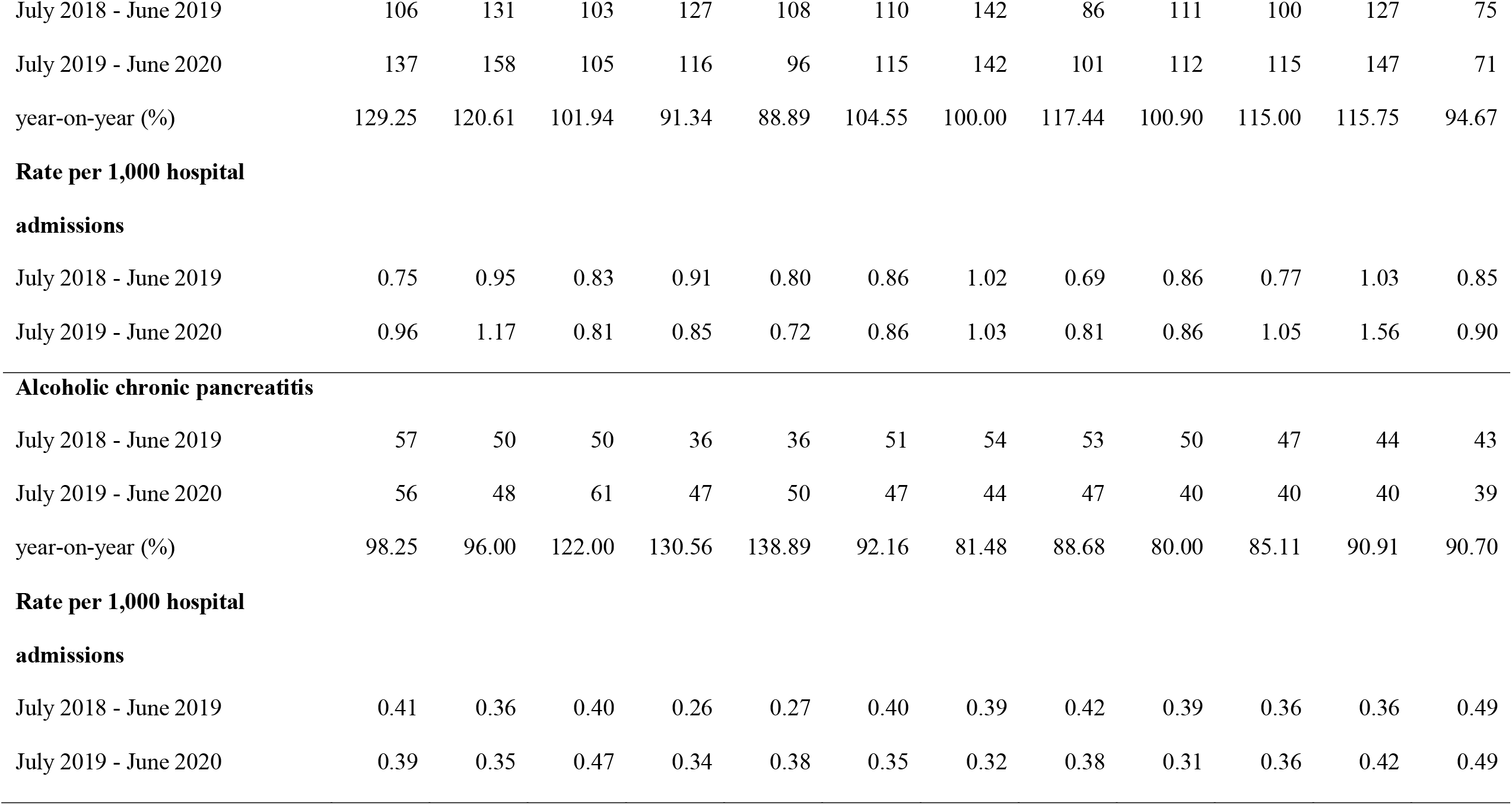
Number of hospital admissions for each disease, and rates (cases/1,000)

Secondary analyses were performed for each diagnosis (alcohol-related liver disease, liver cirrhosis, acute pancreatitis and chronic pancreatitis). The monthly number of hospital admissions, year-on-year and the monthly rates per 1,000 hospital admissions are displayed in Table 2. We display these data together with the predicted regression curves in Figure 2. An increase in the rate of each disease except for chronic pancreatitis was observed after the declaration of emergency for the COVID-19 pandemic by the Japanese government. Compared with acute pancreatitis cases, the monthly rates for cases of alcohol-related liver disease and liver cirrhosis gradually increased after the declaration of emergency. The RR for alcohol-related liver disease was 1.21 (95%CI: 1.08 to 1.35). The RR for liver cirrhosis was 1.21 (95%CI: 1.06 to 1.38). The RR for acute pancreatitis was 1.28 (95%CI: 1.08 to 1.51). The RR for chronic pancreatitis was 1.10 (95%CI: 0.84 to 1.44). The results for the impact of the COVID-19 outbreak were summarized in Table 3.

**Table 3.** Rates of hospital admission for alcohol-related liver disease and pancreatitis during the Outbreak period compared with Pre-outbreak period

| Disease | RR * | Lower 95%CI | Upper 95%CI | P-value |
| --- | --- | --- | --- | --- |
| Alcohol-related liver disease or pancreatitis | 1.22 | 1.12 | 1.33 | <0.001 |
| Alcohol-related liver disease | 1.21 | 1.08 | 1.35 | 0.001 |
| Alcoholic liver cirrhosis | 1.21 | 1.06 | 1.38 | 0.005 |
| Alcoholic acute pancreatitis | 1.28 | 1.08 | 1.51 | 0.004 |
| Alcoholic chronic pancreatitis | 1.10 | 0.84 | 1.44 | 0.467 |

Results adjusted for trend and seasonality
The pandemic period: April 2020 – June 2020
The pre-pandemic period: July 2018 – March 2020
\* Rate ratio: rate of hospital admission for each disease per 1,000 hospital admissions
Abbreviations: CI: Confidence Interval

**Figure 2.**
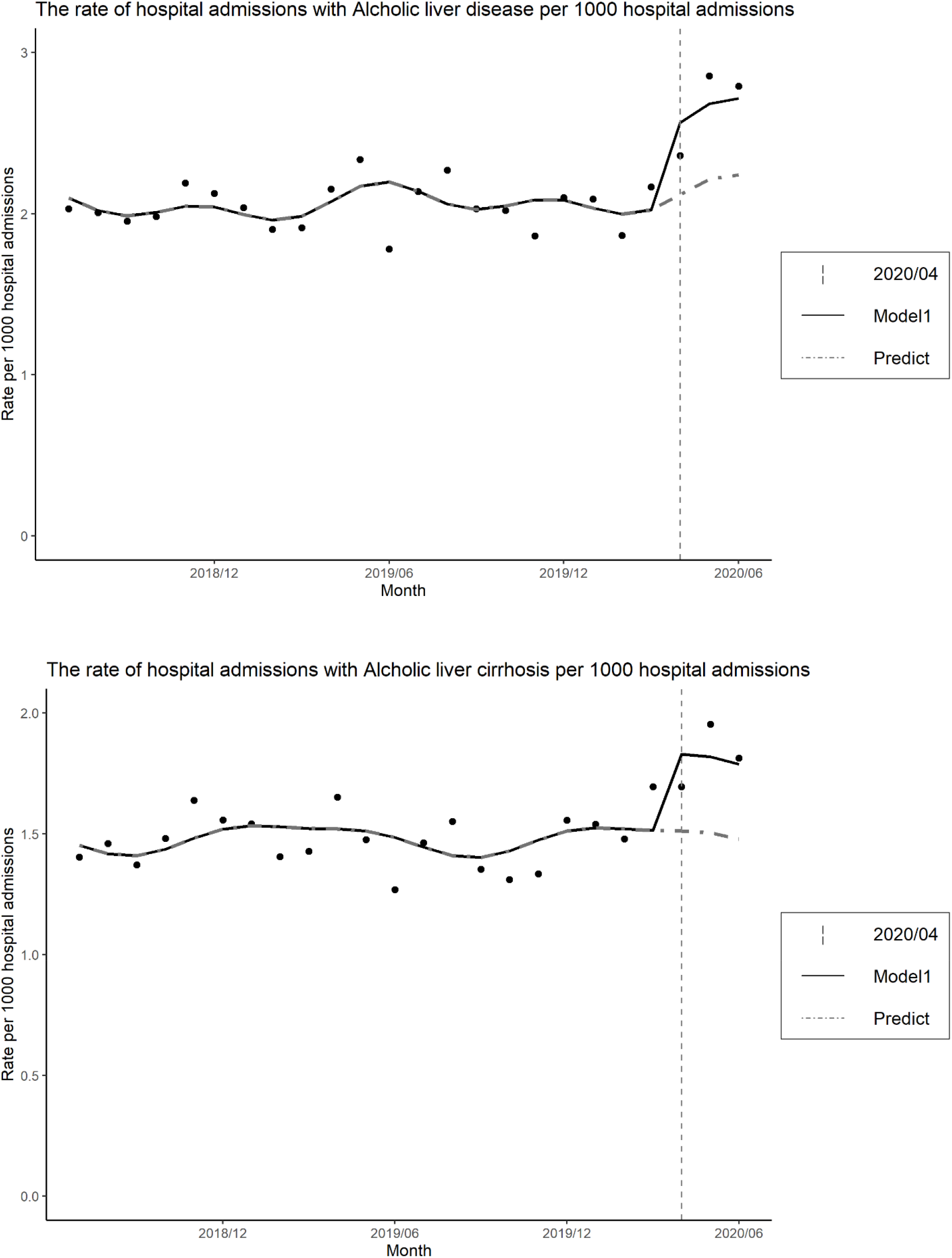
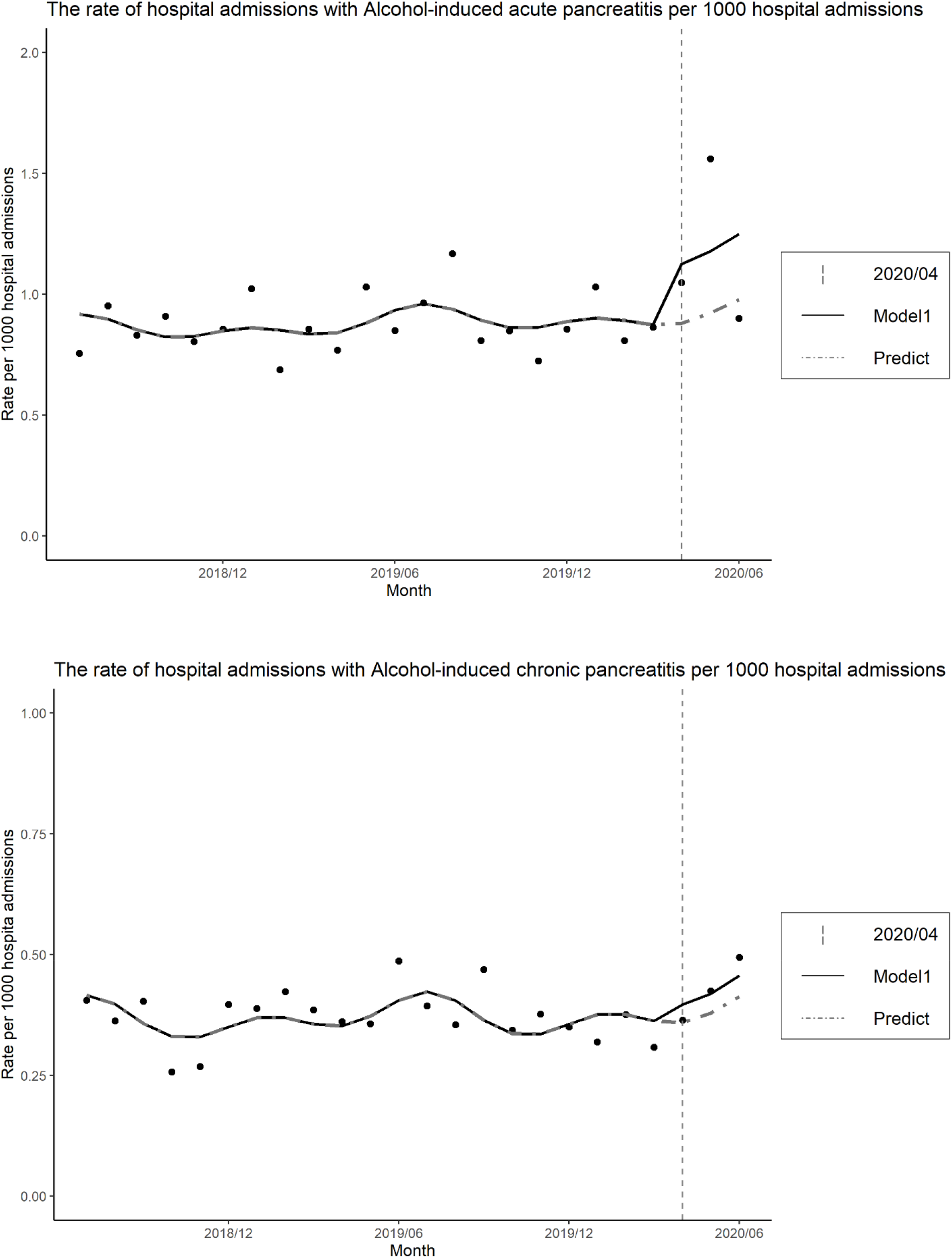
Hospital admissions for each disease during the study period. Model1: Poisson regression model, including trends and seasonality Predict: Counterfactual scenario (if the outbreak did not occur)

The results of the stratified analysis are shown in Supplement Table 3 and 4. They suggested hospital admissions for alcohol-related liver disease or pancreatitis in females might have been more frequent in the COVID-19 outbreak period. Different rates of hospital admissions were not observed for younger and older adults.

## DISCUSSION

In this research, we found that the COVID-19 outbreak was associated with an increase in hospital admissions for alcohol-related liver disease and pancreatitis except for chronic pancreatitis.

The rate of hospital admissions per 1,000 hospital admissions during the COVID-19 outbreak increased by 1.2 times (RR 1.22, 95%CI: 1.12 to 1.33) compared with the pre-pandemic period for cases of alcoholic liver disease or pancreatitis. The COVID-19 outbreak caused about 214.25 excess hospital admissions for alcoholic liver disease or pancreatitis based on predictions from our model.

### Strengths and Limitations

The total case volume was a strength of the study. The total number of hospitalizations was 3,026,389 in 257 hospitals throughout Japan. In contrast, the database had no information about alcohol consumption for each admission case. Therefore, the relationship between individual alcohol consumption and the occurrence of hospital admissions for alcohol-related liver disease or pancreatitis was unclear.

Several studies on alcohol consumption have been published since COVID-19 was first reported [5, 15, 16, 17]. However, few studies have examined the association between the pandemic and alcohol-related physical illnesses, such as liver disease and pancreatitis, as of October 14, 2020, including during the 2003 Severe Acute Respiratory Syndrome (SARS) pandemic. Among more than 800 Hong Kong residents who were exposed to the SARS pandemic in 2003, 4.7% of males and 14.8% of females who were current drinkers reported an increase in drinking one year after the SARS pandemic [18]. In addition, the risk of presenting symptoms of alcohol use disorders three years after the SARS pandemic was about 1.5 times higher for affected individuals, such as health care workers in Beijing who worked in quarantine or high-risk wards, compared with unexposed hospital workers [19].

### Meaning of the study for clinicians and policymakers

Reports of alcohol consumption during the COVID-19 pandemic have varied. Several studies reported that alcohol consumption increased, but other studies reported that alcohol consumption decreased due to fewer opportunities for eating out or shutdown of retailers [5, 15, 16, 17]. In Japan, a survey reported that household expenditure for alcohol after April 2020 increased, while beer companies reported a decrease in sales due to a decrease in sales to the restaurant industry [20, 21]. Our results also suggest that alcoholic cirrhosis accounted for about half of the increase in hospital admissions, which might indicate that alcohol consumption increased in certain high-risk groups rather than in the whole of society. News that alcohol consumption among particular populations such as alcoholics actually increased might support this hypothesis [22–25]. Our exploratory analysis implied that hospital admissions for alcohol-related liver disease or pancreatitis in females might have occurred more frequently compared with males. Pollard et al. reported a significant increase of 0.18 days for heavy drinking (95% CI: 0.04 to 0.32 days) in women in the US, which represents an increase of 41% from a 2019 baseline of 0.44 days [5]. These results suggest that clinicians and policymakers may need to consider medical and policy measures for alcohol misuse in high-risk groups.

### Unanswered questions and future research

Our study could not reveal a relationship between individual alcohol consumption and hospital admissions for alcohol-related liver disease or pancreatitis. Therefore, further research is needed to confirm the association between alcohol consumption and hospital admissions for alcohol-related diseases.

## Conclusions

The COVID-19 outbreak might have increased the hospitalization rate for alcohol-related liver disease and pancreatitis due to an increase in alcohol consumption. Based on the results of this study, clinicians should be aware of the increase in alcohol-related admissions during the pandemic. Moreover, policymakers should keep in mind that measures such as lock-down for the pandemic might increase stress and result in hospitalization for alcohol-related liver disease and pancreatitis.

## Data Availability

The datasets generated during and/or analyzed during the present study are available from the corresponding author on reasonable request.

## Author contributions

Conceptualization: All authors

Methodology: HI, DT and TM

Software: HI, JS, DT, and TM

Validation: HI, JS, DT, TM, SK, and YI

Formal analysis: HI, and JS

Investigation: All authors

Resources: SK and YI

Data Curation: SK

Writing – original draft preparation:

HI Writing – review and editing: All authors

Visualization: HI

Supervision: YI

Project administration: YI

Funding acquisition: YI

## Acknowledgments

The present study was supported by JSPS KAKENHI grant numbers JP19H01075 from the Japan Society for the Promotion of Science and by GAP Fund Program of Kyoto University, Type B (2020) to Yuichi Imanaka.

