## Supplement Table 1, Supplement Table 2, Supplement Table 3, Supplement Table 4 for "The impact of the coronavirus disease 2019 (COVID-19) outbreak on hospital admissions for alcohol-related liver disease and pancreatitis in Japan: a time-series analysis"

**Supplementary material**

**Supplement Table 1. Baseline characteristics of study population (n = 3,026,389)**

|  | Pre-outbreak period  (Jul 2018 to Mar 2020) | | Outbreak period  (Apr 2020 to Jun 2020) | |
| --- | --- | --- | --- | --- |
|  | (n = 2,743,612) |  | (n = 282,777) |  |
| Age |  |  |  |  |
| Median (IQR) | 72 | (59, 81) | 72 | (59, 81) |
| Age category |  |  |  |  |
| 65 years ≥ | 1,884,026 | 68.7% | 191,003 | 67.5% |
| 64 years ≤ | 859,586 | 31.3% | 91,774 | 32.5% |
| Sex |  |  |  |  |
| Males | 1,435,457 | 52.3% | 150,351 | 53.2% |
| Females | 1,308,155 | 47.7% | 132,426 | 46.8% |
| Comorbidity |  |  |  |  |
| ECI, median (IQR) | 0.0 | (0, 5) | 0.0 | (0, 5) |
| Alcohol-related |  |  |  |  |
| Liver disease | 5,623 | 0.20% | 748 | 0.26% |
| Pancreatitis | 3,419 | 0.12% | 452 | 0.16% |
| Length of hospital stay, median (IQR) |  |  |  |  |
| Overall | 8.0 | (4.0, 17.0) | 8.0 | (4.0, 14.0) |
| Liver disease or Pancreatitis | 10.0 | (6.0, 17.0) | 9.0 | (5.0, 15.0) |
| In-hospital mortality | 132,862 | 4.8% | 12,723 | 4.5% |
| Abbreviations: IQR: interquartile range, ECI: Elixhauser Comorbidity Index | | |  |  |

**Supplement Table 2. Number of excess hospital admissions during the Outbreak period (April 2020 to June 2020)**

| Disease | Observed cases^*^ | Counterfactual^†^ | 95%CI | Excess hospital admissions | 95%CI |
| --- | --- | --- | --- | --- | --- |
| Alcohol-related liver disease or pancreatitis | 1,200 | 985.75 | (950.28 to 1,021.22) | 214.25 | (178.78 to 249.72) |
| Alcohol-related liver disease | 748 | 617.45 | (589.34 to 645.55) | 130.55 | (102.45 to 158.66) |
| Alcoholic liver cirrhosis | 513 | 424.04 | (401.24, 446.84) | 88.96 | (66.16 to 111.76) |
| Alcoholic acute pancreatitis | 333 | 260.41 | (242.17, 278.64) | 72.59 | (54.36 to 90.83) |

Excess hospital admissions were defined as the difference in hospital admissions between the actual number of hospital admissions and the predicted number of admissions based on our model.

^*^Observed cases: the actual number of hospital admissions during the Pandemic period

^†^Counterfactual: Predictions based on counterfactual scenario (if the pandemic did not occur)

Abbreviations: CI, Confidence Interval

**Supplement Table 3. Number of hospital admissions for each disease, and rates (cases/1,000) by sex**

|  | Jul | Aug | Sep | Oct | Nov | Dec | Jan | Feb | Mar | Apr | May | Jun |
| --- | --- | --- | --- | --- | --- | --- | --- | --- | --- | --- | --- | --- |
| **Total hospital admissions** |  |  |  |  |  |  |  |  |  |  |  |  |
| **Male** |  |  |  |  |  |  |  |  |  |  |  |  |
| July 2018 - June 2019 | 73,117 | 72,212 | 64,660 | 73,005 | 70,338 | 66,992 | 72,843 | 66,064 | 67,985 | 68,333 | 64,586 | 46,955 |
| July 2019 - June 2020 | 74,078 | 70,558 | 67,836 | 71,017 | 68,885 | 70,189 | 72,113 | 65,567 | 68,124 | 58,196 | 49,454 | 42,701 |
| year-on-year (%) | 101.31 | 97.71 | 104.91 | 97.28 | 97.93 | 104.77 | 99.00 | 99.25 | 100.20 | 85.17 | 76.57 | 90.94 |
| **Female** |  |  |  |  |  |  |  |  |  |  |  |  |
| July 2018 - June 2019 | 67,283 | 65,451 | 59,311 | 66,785 | 63,976 | 61,482 | 66,040 | 59,143 | 61,639 | 61,813 | 58,730 | 41,278 |
| July 2019 - June 2020 | 68,144 | 64,759 | 62,134 | 65,607 | 63,743 | 64,099 | 65,641 | 59,461 | 61,636 | 51,522 | 44,753 | 36,151 |
| year-on-year (%) | 101.28 | 98.94 | 104.76 | 98.24 | 99.64 | 104.26 | 99.40 | 100.54 | 100.00 | 83.35 | 76.20 | 87.58 |
| **Alcohol-related liver disease or pancreatitis** |  |  |  |  |  |  |  |  |  |  |  |  |
| **Male** |  |  |  |  |  |  |  |  |  |  |  |  |
| July 2018 - June 2019 | 377 | 397 | 337 | 373 | 353 | 370 | 411 | 327 | 335 | 371 | 406 | 233 |
| July 2019 - June 2020 | 421 | 435 | 369 | 358 | 320 | 369 | 409 | 323 | 373 | 347 | 381 | 257 |
| year-on-year (%) | 111.67 | 109.57 | 109.50 | 95.98 | 90.65 | 99.73 | 99.51 | 98.78 | 111.34 | 93.53 | 93.84 | 110.30 |
| **Rate per 1,000 hospital admissions** |  |  |  |  |  |  |  |  |  |  |  |  |
| July 2018 - June 2019 | 5.16 | 5.50 | 5.21 | 5.11 | 5.02 | 5.52 | 5.64 | 4.95 | 4.93 | 5.43 | 6.29 | 4.96 |
| July 2019 - June 2020 | 5.68 | 6.17 | 5.44 | 5.04 | 4.65 | 5.26 | 5.67 | 4.93 | 5.48 | 5.96 | 7.70 | 6.02 |
| **Female** |  |  |  |  |  |  |  |  |  |  |  |  |
| July 2018 - June 2019 | 71 | 60 | 58 | 67 | 85 | 64 | 68 | 50 | 74 | 56 | 53 | 42 |
| July 2019 - June 2020 | 76 | 78 | 61 | 81 | 73 | 75 | 65 | 58 | 60 | 67 | 75 | 73 |
| year-on-year (%) | 107.04 | 130.00 | 105.17 | 120.90 | 85.88 | 117.19 | 95.59 | 116.00 | 81.08 | 119.64 | 141.51 | 173.81 |
| **Rate per 1,000 hospital admissions** |  |  |  |  |  |  |  |  |  |  |  |  |
| July 2018 - June 2019 | 1.06 | 0.92 | 0.98 | 1.00 | 1.33 | 1.04 | 1.03 | 0.85 | 1.20 | 0.91 | 0.90 | 1.02 |
| July 2019 - June 2020 | 1.12 | 1.20 | 0.98 | 1.23 | 1.15 | 1.17 | 0.99 | 0.98 | 0.97 | 1.30 | 1.68 | 2.02 |

**Supplement Table 4. Number of hospital admissions for each disease, and rates (cases/1,000) by age (below and above 65 years)**

|  | Jul | Aug | Sep | Oct | Nov | Dec | Jan | Feb | Mar | Apr | May | Jun |
| --- | --- | --- | --- | --- | --- | --- | --- | --- | --- | --- | --- | --- |
| **Total hospital admissions** |  |  |  |  |  |  |  |  |  |  |  |  |
| **Older (65 years ≥)** |  |  |  |  |  |  |  |  |  |  |  |  |
| July 2018 - June 2019 | 95,917 | 92,146 | 84,121 | 96,050 | 92,779 | 88,401 | 97,727 | 86,300 | 89,393 | 89,497 | 84,325 | 57,278 |
| July 2019 - June 2020 | 97,778 | 91,256 | 89,342 | 94,007 | 92,495 | 92,962 | 96,244 | 86,524 | 89,484 | 74,756 | 64,704 | 51,543 |
| year-on-year (%) | 101.94 | 99.03 | 106.21 | 97.87 | 99.69 | 105.16 | 98.48 | 100.26 | 100.10 | 83.53 | 76.73 | 89.99 |
| **Younger (64 years ≤)** |  |  |  |  |  |  |  |  |  |  |  |  |
| July 2018 - June 2019 | 44,483 | 45,517 | 39,850 | 43,740 | 41,535 | 40,073 | 41,156 | 38,907 | 40,231 | 40,649 | 38,991 | 30,955 |
| July 2019 - June 2020 | 44,444 | 44,061 | 40,628 | 42,617 | 40,133 | 41,326 | 41,510 | 38,504 | 40,276 | 34,962 | 29,503 | 27,309 |
| year-on-year (%) | 99.91 | 96.80 | 101.95 | 97.43 | 96.62 | 103.13 | 100.86 | 98.96 | 100.11 | 86.01 | 75.67 | 88.22 |
| **Alcohol-related liver disease or pancreatitis** |  |  |  |  |  |  |  |  |  |  |  |  |
| **Older (65 years ≥)** |  |  |  |  |  |  |  |  |  |  |  |  |
| July 2018 - June 2019 | 139 | 138 | 122 | 149 | 147 | 158 | 165 | 145 | 130 | 122 | 148 | 79 |
| July 2019 - June 2020 | 145 | 140 | 145 | 129 | 144 | 145 | 146 | 140 | 154 | 122 | 125 | 98 |
| year-on-year (%) | 104.32 | 101.45 | 118.85 | 86.58 | 97.96 | 91.77 | 88.48 | 96.55 | 118.46 | 100.00 | 84.46 | 124.05 |
| **Rate per 1,000 hospital admissions** |  |  |  |  |  |  |  |  |  |  |  |  |
| July 2018 - June 2019 | 1.45 | 1.50 | 1.45 | 1.55 | 1.58 | 1.79 | 1.69 | 1.68 | 1.45 | 1.36 | 1.76 | 1.38 |
| July 2019 - June 2020 | 1.48 | 1.53 | 1.62 | 1.37 | 1.56 | 1.56 | 1.52 | 1.62 | 1.72 | 1.63 | 1.93 | 1.90 |
| **Younger (64 years ≤)** |  |  |  |  |  |  |  |  |  |  |  |  |
| July 2018 - June 2019 | 309 | 319 | 273 | 291 | 291 | 276 | 314 | 232 | 279 | 305 | 311 | 196 |
| July 2019 - June 2020 | 352 | 373 | 285 | 310 | 249 | 299 | 328 | 241 | 279 | 292 | 331 | 232 |
| year-on-year (%) | 113.92 | 116.93 | 104.40 | 106.53 | 85.57 | 108.33 | 104.46 | 103.88 | 100.00 | 95.74 | 106.43 | 118.37 |
| **Rate per 1,000 hospital admissions** |  |  |  |  |  |  |  |  |  |  |  |  |
| July 2018 - June 2019 | 6.95 | 7.01 | 6.85 | 6.65 | 7.01 | 6.89 | 7.63 | 5.96 | 6.93 | 7.50 | 7.98 | 6.33 |
| July 2019 - June 2020 | 7.92 | 8.47 | 7.01 | 7.27 | 6.20 | 7.24 | 7.90 | 6.26 | 6.93 | 8.35 | 11.22 | 8.50 |
